# Carriage of ESBL-producing Enterobacterales in wastewater treatment plant workers and surrounding residents - The AWARE Study

**DOI:** 10.1101/2021.06.25.21259524

**Authors:** Daloha Rodríguez-Molina, Fanny Berglund, Hetty Blaak, Marcela Popa, Carl-Fredrik Flach, Merel Kemper, Luminita Marutescu, Gratiela Pircalabioru, Beate Spießberger, Tobias Weinmann, Laura Wengenroth, Mariana Carmen Chifiriuc, D. G. Joakim Larsson, Katja Radon, Dennis Nowak, Andreas Wieser, Ana Maria de Roda Husman, Heike Schmitt

## Abstract

**Purpose:** To investigate whether wastewater treatment plant (WWTP) workers and residents living in close proximity to a WWTP have elevated carriage rates of ESBL-producing Enterobacterales, as compared to the general population.

**Methods:** From 2018 to 2020, we carried out a cross-sectional study in Germany, the Netherlands, and Romania among WWTP workers (N=344), nearby residents (living ≤ 300 meters away from WWTPs; N=431) and distant residents (living ≥ 1000 meters away = reference group; N=1165). We collected information on potential confounders via questionnaire. Culture of participants’ stool samples was performed with ChromID®-ESBL agar plates and species identification with MALDI-TOF-MS. We used logistic regression to estimate the odds ratio (OR) for carrying ESBL-producing *E. coli* (ESBL-EC). Sensitivity analyses included stratification by country and interaction models using country as secondary exposure.

**Results:** Prevalence of ESBL-EC was 11% (workers), 29% (nearby residents), and 7% (distant residents), and higher in Romania (28%) than in Germany (7%) and the Netherlands (6%). Models stratified by country showed that within the Romanian population, WWTP workers are about twice as likely (aOR = 2.34, 95% CI: 1.22-4.5) and nearby residents about three times as likely (aOR = 3.17, 95% CI: 1.8-5.59) to be ESBL-EC carriers, when compared with distant residents.

**Conclusions:** In stratified analyses by country, we found an increased risk for carriage of ESBL-EC in Romanian workers and nearby residents. This effect was higher for nearby residents than for workers, which suggests that, for nearby residents, factors other than the local WWTP could contribute to the increased carriage.

## Introduction

Antibiotic resistance (AR) is currently one of the most important threats to public health and clinical medicine. In some regions, current AR rates are alarmingly high, with 58.4% of *Escherichia coli* (*E. coli*) isolates reported in 2018 to the European Antimicrobial Resistance Surveillance Network being resistant to at least one antibiotic group under surveillance (i.e. aminopenicillins, fluoroquinolones, third-generation cephalosporins, aminoglycosides and carbapenems) [1]. This is partly due to the use, overuse, and misuse of antibiotics by healthcare professionals and patients, but also in animal husbandry and agriculture [2–6]. Antibiotic resistant bacteria (ARB) can be introduced into the environment by different routes [7], including wastewater from the general human population [8–15]. These residual waters arrive and are collected at municipal wastewater treatment plants (WWTPs). Enteric ARB such as *E. coli*, as well as *Klebsiella spp*., *Enterobacter spp*., *Serratia spp*., and *Citrobacter spp*. (KESC) have been found in water [16–22] and air [23–25] samples from WWTPs. Moreover, the WWTPs effluents can discharge ARB into nearby water bodies because eliminating ARB is not part of current wastewater treatment processes, which focus instead on reducing nutrient loads and pathogens to the receiving surface water. While some studies have reported either no changes in relative abundances of ARB [26] or a decrease in absolute and relative abundance of ARGs [27–29], other studies have reported an increased relative prevalence of ARB after wastewater treatment processes, in comparison to the untreated wastewater entering the plant [16,17,22,30–38]. These aspects make WWTPs potential transmission hubs for the spread of ARB into the environment [39].

It has been proposed that ARB could be transmitted to humans by the air or wastewater at the WWTPs through different exposure routes including ingestion of droplets, hand-to-mouth contact, or inhalation of aerosols [21–24]. Further, an increased prevalence of gastrointestinal and respiratory diseases [40], as well as high levels of antibodies against bacteria, viruses, and parasites in WWTP workers, suggests an increased exposure to these pathogens [41–43]. Under this scenario, and extending this idea to AR, WWTP workers would be at a high risk of exposure to ARB. Furthermore, and considering that extended-spectrum betalactamase (ESBL)-producing *E. coli* (ESBL-EC) can be found up to 150 meters both up- and downwind away from animal farms [44], nearby residents living in close proximity to WWTPs could also be highly exposed to these ARB. However, to our knowledge, no large-scale study has yet been carried out in humans potentially at risk of carriage of antibiotic resistant Enterobacterales working at or living close to WWTPs. Such studies are critical to aid our current understanding of the exposure status of humans working at or living around WWTPs, and to devise preventive strategies and interventions to reduce this potential exposure.

Therefore, in the present study we aimed at investigating whether WWTP workers and residents living in close proximity to a WWTP have elevated carriage rates of ESBL-producing Enterobacterales, as compared to the general population. Our hypothesis is that the risk of carrying ESBL-producing Enterobacterales increases with proximity to the WWTP.

## Materials & Methods

### Study design and population

The project “Antibiotic Resistance in Wastewater: Transmission Risks for Employees and Residents around Wastewater Treatment Plants (AWARE)” is a cross-sectional study, with data collection carried out from September 2018 to March 2020 in three European countries with different background prevalences for AR: Germany, the Netherlands, and Romania. A thorough description of the study methodology can be found elsewhere [45]. Briefly, our target population consisted of two exposed groups working at or living in close proximity to wastewater treatment plants (WWTP workers and nearby residents) and one unexposed population of distant residents. Nearby residents were defined as living within a 300-meter radius from a WWTP, while distant residents were defined as living more than 1000 meters away from a WWTP. Data on nearby residents was only collected in Germany and Romania, while data on WWTP workers and distant residents was collected in all three countries. The process of recruiting participants per country is described as follows.

#### Germany

We generated a sampling frame of WWTPs and ranked them in descending order based on number of employed workers and of estimated nearby residents in their vicinity to maximize the chances of achieving the minimum sample size for these two exposed groups. Out of 18 eligible WWTPs with the largest number of employed workers and nearby residents, eight were interested in participating and were thus invited into the study. Of these eight plants, six were willing to participate, of which one had too few workers and was thus not eligible, one could not participate anymore because of the situation regarding COVID-19 in early 2020, and one was selected as a pilot phase plant because it had a lower number of workers and nearby residents (Fig. 1). The remaining three plants were enrolled in full participation.

**Fig. 1:**
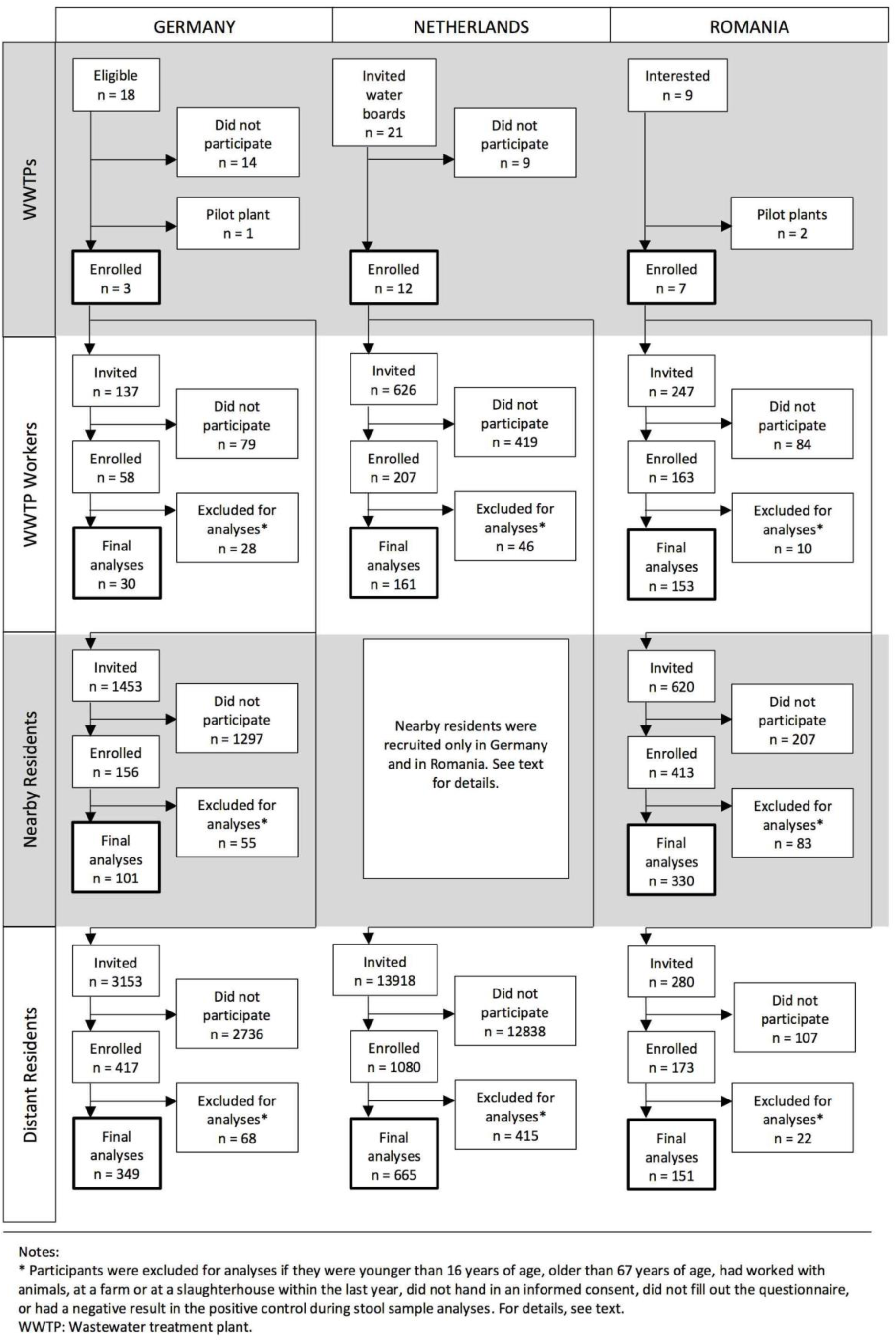
Flow diagram of the recruitment process, AWARE Study, 2021

After a pilot phase examining the feasibility of the study methods, a total of 137 workers employed at three WWTPs in Southern Germany were invited to participate in our study (response 22%). For nearby and distant residents of each of these three WWTPs, postal addresses were obtained from the local civil registries whenever possible, and all individuals living at each household were invited to participate in our study via postal service. In study locations where this was not possible, we generated a sampling frame of addresses within the specified distances to the WWTP for nearby and distant residents using Google Maps™, and went door-to-door delivering invitation letters to mailboxes. In addition to the invitation letter, two reminders were sent to non-responders. In parallel, local newspapers published an article about the project on the same week that the participants received the invitation letter. We also carried out a recruitment campaign via Facebook, targeting potential participants within the desired age range and located at the study sites. All participants who successfully completed the study were eligible for a raffle of shopping vouchers with a total value of 1,500 EUR. In total, we invited 1453 nearby residents within the eligible age range (response 6.95%) and 3153 distant residents (response 11%).

#### The Netherlands

In the Netherlands, WWTPs are managed by regional water authorities called waterboards. Our unit of recruitment for the Netherlands was therefore the waterboard and not the WWTP. Of a total of 21 waterboards across the whole country, 12 were interested in participating in the study. Overall, 626 WWTP workers were invited to participate using a combination of WWTP visits for presenting the study plus invitations by e-mail in ten out of these twelve WWTPs, and using only e-mail invitations in the remaining two plants (response 26%). We did not carry out data collection for residents living in close proximity to WWTPs in the Netherlands. For distant residents, general practitioners (GP) practices located 2 to 5 km away from the selected WWTPs were identified and these GPs were invited to cooperate with us as their practices served as a collection and preservation point for stool samples. Using ArcGis[46], we then identified all postal addresses within a 500-meter radius from the cooperating GP practices, and then, using the Dutch Personal Records Database we randomly retrieved the contacting information of potential participants living in 300-500 addresses surrounding each GP practice. A total number of 13,918 individuals living at these addresses received an invitation letter per postal service, of which 1,080 responded to the invitation (recruitment response 7.8%). Of these 13,918 invited people, 10,448 individuals were between the age of 16 and 67 years old and thus eligible by age (response among eligible individuals 6.4%). All participants completing the study received a gift card worth 20 EUR.

#### Romania

WWTP operators were recruited through a formal letter containing information about the project and an invitation to join the study. Nine plants were invited, of which two were pilot plants, and all of them were ultimately enrolled in the study. WWTP workers from participating plants were contacted by their respective operators and invited to participate. A total number of 247 workers were reached (response 62%). Nearby and distant residents were invited to participate using the door-to-door approach. Further, potential participants in public places like streets, parks, and markets in the vicinity of WWTPs were also addressed orally and invited to participate, given that they were eligible. In total, we contacted 620 nearby and 280 distant residents within the eligible age range (response 53% and 54%).

### Data collection

#### Exposure of interest

We consider ingestion of droplets, hand-to-mouth contact, or inhalation of aerosols the main exposure routes for WWTP workers. Nearby residents would be exposed through inhalation of aerosols. Therefore, we used the variable participation group (WWTP worker, nearby resident, distant resident) as a proxy variable for the exposure. We defined WWTP workers as the highest exposed group followed by nearby residents as the second most-exposed group, while distant residents served as an unexposed comparison group. Nearby residents were defined as persons living fewer than 300 meters away from the WWTP. Distant residents were defined as persons living further than 1000 meters away from any WWTPs.

#### Outcome of interest

The main outcome of interest was the presence of ESBL-EC in stool samples, reported binarily (positive/negative). A secondary outcome of interest was the presence of bacteria from the *Klebsiella, Enterobacter, Citrobacter*, and *Serratia* (KESC) group in stool samples, also reported binarily (positive/negative). In Germany and Romania, only participants who successfully filled in the study questionnaire were sent a stool sample kit. In the Netherlands, enrolled participants were required to hand in a stool sample before receiving a link to fill in the online questionnaire. Nearby and distant residents received a stool sample collection kit by postal service, whereas workers received it at their workplace. Each participant was asked to record the date and time of stool sample collection, maintain the sample refrigerated (temperature ranging from 2 °C to 8 °C), and bring it to the closest collection point (WWTPs or main train station in Germany, WWTPs or GP offices in the Netherlands, home visits in Romania). Samples were transported to the laboratory in cooling boxes within 24 hours after sampling, where they were stored at 4 °C, and processed within 24 - 48 hours after sampling.

At the local laboratories in Germany, the Netherlands and Romania, all the stool samples were inoculated directly onto the following culture media: ChromID® ESBL (for ESBL-EC), TBX (in the Netherlands and Romania) or MacConkey (in Germany) (for *E. coli*), and incubated at 36 °C ± 1 °C for 24 - 48 h. In case of positive results, 2 separate isolates belonging to the ESBL-EC phenotype were collected from the ChromID® ESBL plate, screened for antibiotic resistance and identified by MALDI-TOF MS (Matrix Assisted Laser Desorption Ionization-Time of Flight Mass Spectrometry). Participants with a negative stool culture on TBX/MacConkey were excluded from further analyses.

#### Confounding variables

Information on confounding variables was obtained from eligible individuals through an online questionnaire exploring sociodemographic characteristics, work history including contact with animals during farming or slaughterhouse activities, contact with patients or human tissues at work, international travels, use of antibiotics, hospital visits, and health condition (personal history of surgery, hospitalizations, chronic diseases, antibiotic and antacid intake, diarrhea, respiratory health, and self-reported health status), all in the past 12 months [45].

Educational level was asked using the educational structure of each country and then dichotomized using the International Standard Classification of Education (ISCED) [47–49] into low (0 - 2 ISCED points, i.e. pre-primary education to lower secondary education) and high (more than 2 ISCED points, i.e. upper secondary education to Doctoral or equivalent).

Work with patients or human tissues was constructed by merging the information of two separate survey questions: “In your current job, how often have you typically had direct interaction or contact with patients within in the last 12 months?” and “How often have you worked with human tissue, blood, body fluids (urine, feces, vomit, sputum, saliva) or primary cell lines within the last 12 months?” Each question could be answered with a frequency scale (never, rarely, sometimes, often, always). If the participant had answered rarely, sometimes, often or always in either of the two questions, a “yes” was assigned. Else, a “no” was assigned. Use of antibiotics was assessed with the question “Have you taken an antibiotic within the last 12 months?” to which possible answers were “Yes,” “No,” and “Do not know.” Participants answering “Do not know” were assigned into the “No” category.

When asked about international travel, participants were asked to provide information about the region where they had been in the past year: Europe, Asia, North Africa, Sub-Saharan Africa, North America, Central America and Mexico, South America, and Australia and Oceania. For each of these regions, participants could state the frequency of travel within the last year: never, once, 2 to 3 times, more than 3 times, I don’t know. Additionally, if the participant reported travels to Europe, they were asked about travels to specific European countries with a high background prevalence of ESBL-EC: Italy, Slovenia, Bulgaria, and Greece (yes/no). Travels to high-risk areas for ESBL was defined as reporting travels to at least one of the following areas or countries within the past year: Asia, North Africa, Sub-Saharan Africa, Central America and Mexico, South America, Italy, Slovenia, Bulgaria, and Greece.

### Statistical analyses

To present summary statistics for the descriptive characteristics of the study population, numerical variables (i.e. age) were assessed visually for normality using histograms and are presented as mean ± standard deviation if normally distributed or as median ± inter-quartile range if non-normally distributed. Categorical variables are presented using absolute and relative frequencies. Either chi-square of Fisher’s exact test was used for bivariate hypothesis testing of categorical variables, depending on cell counts.

We assume that the missing values in the outcome of interest are missing at random because it is highly unlikely that participants would know their personal status of ESBL-EC in stools beforehand. We therefore proceeded to simulate missing values for this outcome and other variables of interest where the missingness mechanism was at random or completely at random by using multiple imputation with chained equations [50]. With twenty iterations per dataset, we generated a total of ten imputed datasets, from which we estimated regression models whose estimates were then pooled and reported. Because of the differences in participation response across countries, we weighted our study population using inverse probability of sampling weights [51]. Weights were defined as the inverse of the participation response per country and per participation group.

The direct causal effect of participation group (WWTP worker, nearby resident, distant resident) as a proxy for exposure routes (ingestion of droplets, hand-to-mouth contact, or inhalation of aerosols) in and around the local WWTP on the presence of ESBL-EC in participants’ stool samples (no/yes) was estimated using logistic regression models. We present unweighted crude and adjusted estimates, weighted crude and adjusted estimates, and their corresponding 95% confidence intervals in graphical form. Sensitivity analyses included models stratified by country, an interaction model with country as a secondary exposure, and models stratified by participation group.

Variable selection for the models was done using a combination of experts’ opinion from within the AWARE consortium, evidence in the current literature, and the use of Directed Acyclic Graphs (DAGs) [52,53] (Fig. 2). All analyses were done in R version 3.5.0 and up [54] using the following R packages: epiR [55], mice [56], mitml[57], mitools[58], and survey[59,60].

**Fig. 2:**
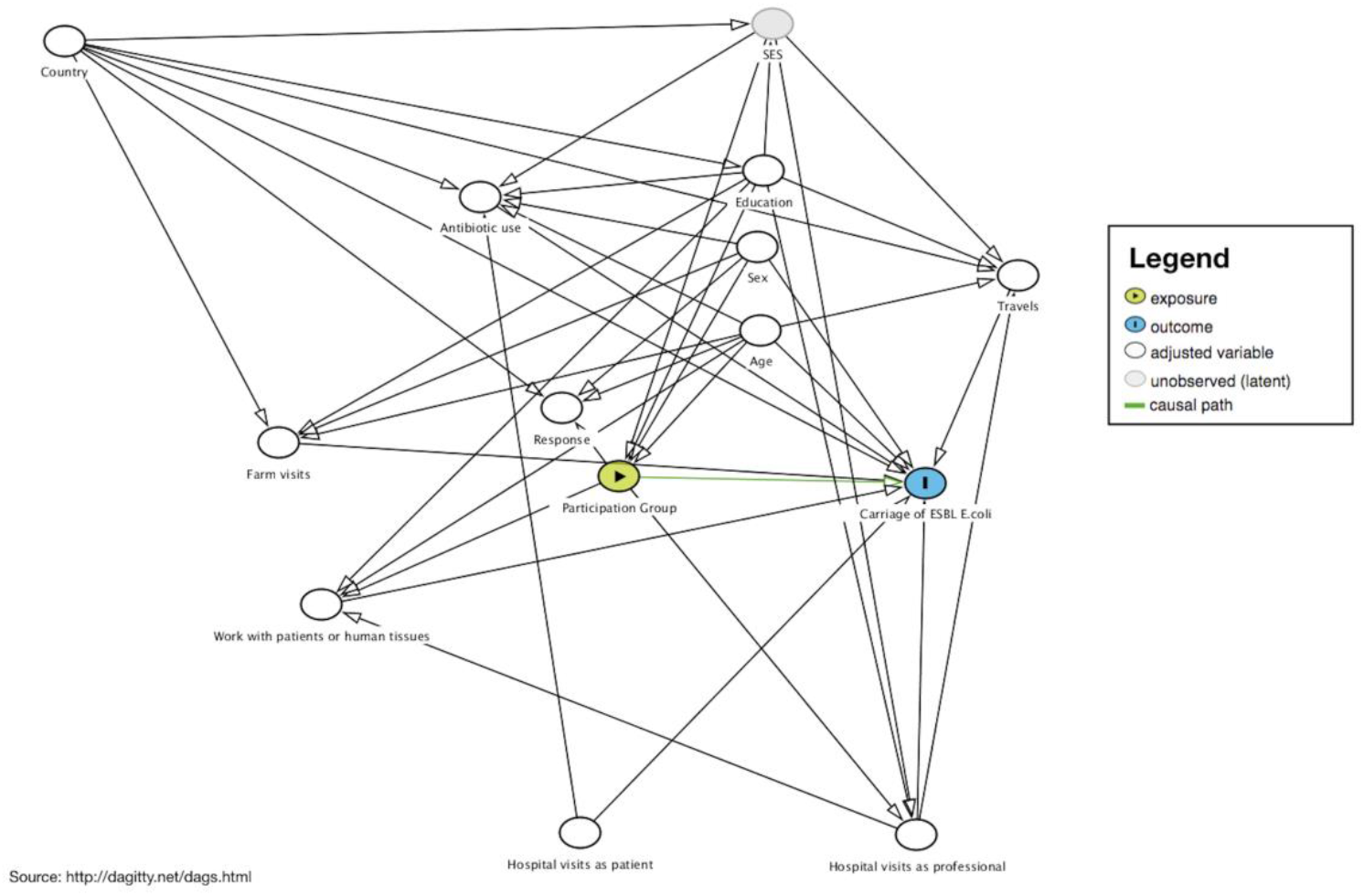
Directed Acyclic Graph (DAG) for the direct effect of participation group (wastewater treatment plant -WWTP-worker, nearby resident, distant resident) as a proxy for exposure routes (ingestion of droplets, hand-to-mouth contact, or inhalation of aerosols) in and around the local WWTP on the presence of ESBL-producing E. coli in stool samples, AWARE Study, 2021

## Results

### Descriptive characteristics of the study population

A total of 1940 participants across the three countries were eligible for analyses, with 25% of participants from Germany (n = 480), 43% from the Netherlands (n = 826), and 33% from Romania (n = 634, Table 1). The majority of the population was middle-aged (median age 49 years, IQR 36-58), female (52%), and highly educated (64%). Across the three countries, WWTP workers were mostly men and the majority reported contact with human tissues, which we attribute to the presence of human feces in wastewater.

**Table 1:** Descriptive characteristics of the studied population by country and participation group, n = 1940, AWARE Study, 2021

| Variable | Missings | Overall | Germany |  |  |  |  | The Netherlands<sup>a</sup> |  |  |  | Romania |  |  |  |  |
| --- | --- | --- | --- | --- | --- | --- | --- | --- | --- | --- | --- | --- | --- | --- | --- | --- |
|  |  |  | Overall | WWTP worker | Nearby resident<sup>c</sup> | Distant resident<sup>b</sup> | p | Overall | WWTP worker | Distant resident<sup>b</sup> | p | Overall | WWTP worker | Nearby resident<sup>c</sup> | Distant resident<sup>b</sup> | p |
| n |  | 1940 | 480 | 30 | 101 | 349 |  | 826 | 161 | 665 |  | 634 | 153 | 330 | 151 |  |
| Age, years (median [IQR]) | 0 | 49<br>[36, 58] | 47<br>[35, 57] | 52<br>[44, 55] | 48<br>[35, 58] | 45<br>[34, 56] | 0.161 | 54<br>[40, 61] | 54<br>[45, 59] | 55<br>[39, 61] | 0.710 | 43<br>[34, 53] | 49<br>[41, 53] | 41<br>[32, 54] | 40<br>[33, 50] | <0.001 |
| Sex, n (%) = Male | 4 | 938<br>(48) | 211<br>(44) | 24 (80) | 51 (50) | 136 (39) | <0.001 | 403<br>(49) | 150 (93) | 253 (38) | <0.001 | 324<br>(51) | 114 (75) | 140 (42) | 70 (47) | <0.001 |
| Highest educational level obtained, n (%) = High<sup>d</sup> | 8 | 1228<br>(64) | 307<br>(64) | 9 (30) | 47 (47) | 251 (72) | <0.001 | 426<br>(52) | 41 (25) | 385 (58) | <0.001 | 495<br>(79) | 144 (97) | 214 (65) | 137 (91) | <0.001 |
| Work with patients or human tissues in the past year, n (%) = Yes<sup>e</sup> | 43 | 605<br>(32) | 171<br>(36) | 19 (68) | 32 (32) | 120 (35) | 0.001 | 321<br>(39) | 96 (62) | 225 (34) | <0.001 | 113<br>(18) | 25 (18) | 50 (15) | 38 (26) | 0.025 |
| Hospital visits as a patient in the past year, n (%) = Yes | 2 | 172 (9) | 74 (15) | 2 (7) | 18 (18) | 54 (16) | 0.332 | 42 (5) | 2 (1) | 40 (6) | 0.024 | 56 (9) | 9 (6) | 38 (12) | 9 (6) | 0.046 |
| Hospital visits as a professional in the past year, n (%) = Yes | 2 | 59 (3) | 31 (6) | 0 (0) | 6 (6) | 25 (7) | 0.299 | 14 (2) | 0 (0) | 14 (2) | 0.131 | 14 (2) | 0 (0) | 7 (2) | 7 (5) | 0.023 |
| Use of antibiotics in the past year, n (%) = Yes | 4 | 454<br>(23) | 147<br>(31) | 7 (23) | 27 (27) | 113 (32) | 0.372 | 147<br>(18) | 17 (11) | 130 (20) | 0.010 | 160<br>(25) | 31 (21) | 83 (25) | 46 (30) | 0.156 |
| Farm visits in the past year, n (%) = Yes | 9 | 181 (9) | 85 (18) | 10 (33) | 14 (14) | 61 (17) | 0.050 | 79 (10) | 25 (16) | 54 (8) | 0.005 | 17 (3) | 2 (1) | 7 (2) | 8 (5) | 0.067 |
| Travel to high risk areas for AR in the past year, n (%) = Yes<sup>f</sup> | 18 | 658<br>(34) | 241<br>(51) | 13 (43) | 42 (42) | 186 (54) | 0.083 | 291<br>(36) | 52 (33) | 239 (36) | 0.501 | 126<br>(20) | 45 (30) | 32 (10) | 49 (33) | <0.001 |
| Carriage of ESBL-producing E. coli, n (%) = Positive | 163 | 236<br>(13) | 26 (7) | 0 (0) | 5 (6) | 21 (8) | 0.218 | 47 (6) | 7 (4) | 40 (6) | 0.532 | 163<br>(28) | 27 (23) | 118 (36) | 18 (12) | <0.001 |
| Carriage of ESBL-producing KESC bacteria, n (%) = Positive | 163 | 67 (4) | 4 (1) | 0 (0) | 1 (1) | 3 (1) | 0.845 | 4 (0) | 3 (2) | 1 (0) | 0.029 | 59 (10) | 12 (10) | 35 (11) | 12 (8) | 0.740 |

Notes:
<sup>a</sup>No data from nearby residents were collected in the Netherlands.
<sup>b</sup>Distant residents live at least 1000 m away from a WWTP
<sup>c</sup>Nearby residents live within a 300 m radius from a WWTP.
<sup>d</sup>Educational level according to the International Standard Classification of Education (ISCED): Low = ISCED 0-2 (Pre-primary education to Lower secondary education), High = ISCED ≥3 (Upper secondary education to Doctoral or equivalent).
<sup>e</sup>Work with patients or human tissues in the past year: Includes self-reported contact with patients at work and with human tissues (e.g. blood, urine, sputum, feces, vomit, saliva, or primary cell lines).
<sup>f</sup>Travel to high risk areas for AR in the past year: Includes travels to North Africa, Sub-Saharan Africa, Asia, Central and South America, as well as the European countries Italy, Greece, Bulgaria and Slovenia.
ESBL: Extended-Spectrum Beta-Lactamases.
AR: Antibiotic Resistance.

In Germany, approximately two-thirds of the WWTP workers reported working with human tissues (68%) in contrast to nearby and distant residents, where approximately a third of each group reported this type of contact at work (32% and 35%, p 0.0015). Distant residents from Germany were more highly educated than nearby residents, and these in turn more than WWTP workers (72%, 47%, and 30%, p <0.001).

In the Netherlands, fewer WWTP workers reported using antibiotics in the past year in comparison to the distant residents (11% vs. 20%, p = 0.01) and visiting hospitals as a patient (1.2% vs. 6.0%, p = 0.02). More WWTP workers reported visiting farms than distant residents (16% vs. 8.2%, p = 0.005).

In Romania, workers were, on average, older (median age among workers 49 [41, 53] vs. median age among distant residents 40 [33, 50] in distant residents) and better educated (97% vs. 91) than distant residents. Also, in comparison to distant residents, nearby residents had a lower level of education (65% vs. 91%) and traveled less to high risk areas for AR (10% vs. 33%).

### Carrier status for ESBL-producing Enterobacterales

The overall prevalence of ESBL-EC across the three countries was 13%, with the highest prevalence observed in the Romanian population (28%). The prevalence of ESBL-producing bacteria of the KESC group across countries was 3.8%, with the highest value observed also in Romania (10%).

In Germany, ESBL-EC were not detected in stools of any of the workers (n = 30), but among 8.4% of distant residents and 5.7% of nearby residents. In the Netherlands, carriage of ESBL-EC was similar in WWTP workers (4.4%) and distant residents (6.0%) (p = 0.53). In Romania, the prevalence of ESBL-EC was 23% among workers, 36% among nearby residents, and 12% among distant residents (p <0.001).

Because the prevalence for KESC bacteria was relatively low and thus limiting the statistical power of our inferential analyses, we decided to focus only on the primary outcome: ESBL-EC. The effect of participation group (WWTP worker, nearby or distant resident) on the carriage of ESBL-EC varied by country (Online Resource Table 1). Overall, the proportion of WWTP workers and nearby residents with a positive stool sample for ESBL-EC was higher than that of distant residents (11% and 29% vs. 7.5%, p <0.001). This result was driven by the Romanian population (23% and 36% vs. 12%, p <0.001), while there were no statistically significant differences between participation groups in the proportions of positive ESBL-EC carriers either in Germany (0.0% and 5.7% vs. 8.4%, p = 0.22) or in the Netherlands (4.4% vs. 6.0%, p = 0.53).

### Statistical models

Across the three countries, the unweighted crude odds ratio for the carriage of ESBL-EC among WWTP workers was 1.71 (95% CI: 1.12-2.61). Among nearby residents, it was 4.95 (95% confidence interval, CI: 3.63-6.73), compared to the unexposed group (Fig. 3). These unweighted estimates changed to 1.17 (95% CI: 0.74-1.86) for WWTP workers and 2.24 (95% CI: 1.5-3.37) for nearby residents upon adjustment for age, sex, education, country, travels to high risk areas for AR, working with human tissues, antibiotic use, farm visits, hospital visits as patients, and hospital visits as a professional. After applying inverse probability of sampling weights for the response in each country and in each participation group, crude estimates changed to 1.28 (95% CI: 0.82-2) among workers and to 2.46 (95% CI: 1.65-3.69) among nearby residents, while the adjusted estimates changed to 0.76 (95% CI: 0.44-1.29) and 1.47 (95% CI: 0.83-2.59), respectively.

**Fig. 3:**
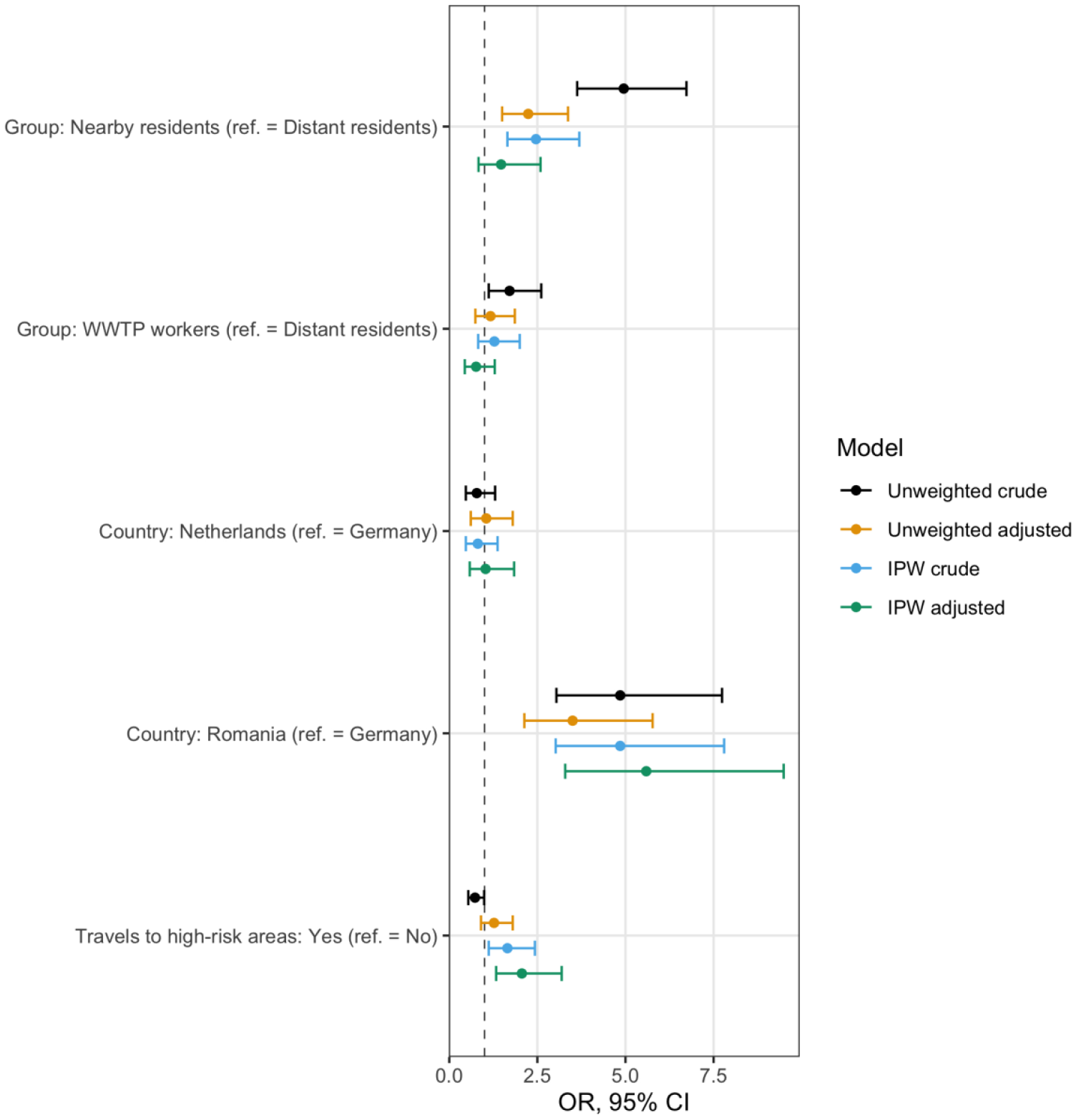
Comparison of models estimating the effect of participation group (wastewater treatment plant - WWTP- worker, nearby resident, distant resident) as a proxy for exposure routes (ingestion of droplets, hand-to-mouth contact, or inhalation of aerosols) in and around the local WWTP on the presence of ESBL-producing E. coli in stool samples, AWARE Study, 2021. Models adjusted for age, sex, education, country, travels to high risk areas, working with human tissues, antibiotic use, farm visits, hospital visits as patient and hospital visits as a professional. IPW: Inverse Probability Weighted model. ref. = Reference level. Travel to high risk areas for AR in the past year includes travels to North Africa, Sub-Saharan Africa, Asia, Central and South America, as well as the European countries Italy, Greece, Bulgaria and Slovenia. Crude: Model with only the given variable, ignoring potential covariates. Adjusted: Model with the given variable, including all potential covariates in the exposure-outcome relation. Unweighted: Model without applying inverse probability weights (IPW). Weighted: Model applying inverse probability weights (IPW). See text for details

Although we could not estimate an effect of exposure within the German and the Dutch subpopulations (Table 2), models stratified by country showed that, within the Romanian population, WWTP workers were about twice as likely (adjusted OR, aOR = 2.34, 95% CI: 1.22-4.5) and nearby residents about three times as likely (aOR = 3.17, 95% CI: 1.8-5.59) to be ESBL-EC carriers, when compared with distant residents.

**Table 2:**
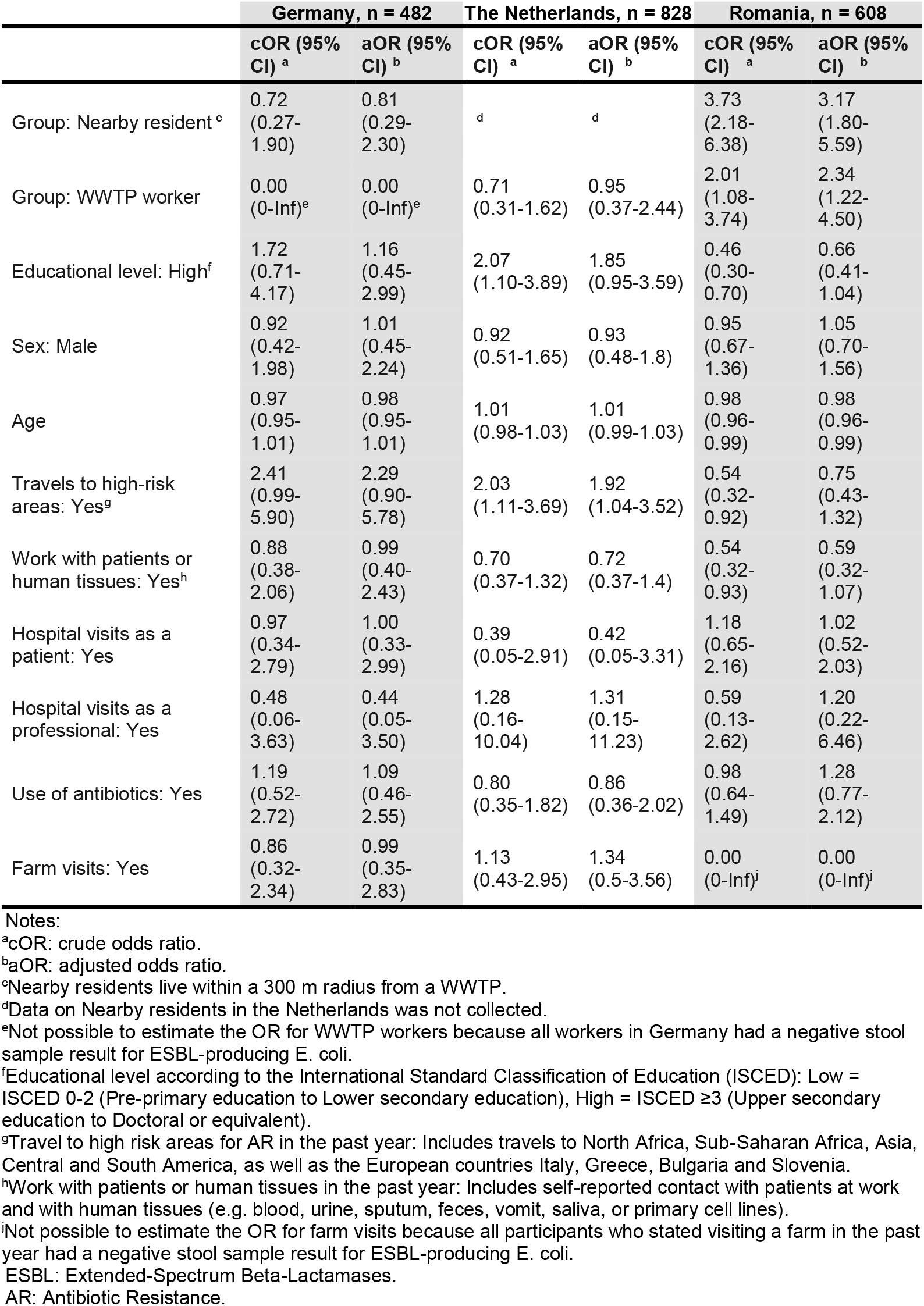
Unweighted models for the carriage of ESBL-producing E. coli, stratified by country, n = 1940, AWARE Study, 2021

|  | Germany, *n* = 482 |  | The Netherlands, *n* = 828 |  | Romania, *n* = 608 |  |
| --- | --- | --- | --- | --- | --- | --- |
|  | cOR (95% CI)<sup>a</sup> | aOR (95% CI)<sup>b</sup> | cOR (95% CI)<sup>a</sup> | aOR (95% CI)<sup>b</sup> | cOR (95% CI)<sup>a</sup> | aOR (95% CI)<sup>b</sup> |
| Group: Nearby resident<sup>c</sup> | 0.72<br>(0.27-1.90) | 0.81<br>(0.29-2.30) | <sup>d</sup> | <sup>d</sup> | 3.73<br>(2.18-6.38) | 3.17<br>(1.80-5.59) |
| Group: WWTP worker | 0.00<br>(0-Inf)<sup>e</sup> | 0.00<br>(0-Inf)<sup>e</sup> | 0.71<br>(0.31-1.62) | 0.95<br>(0.37-2.44) | 2.01<br>(1.08-3.74) | 2.34<br>(1.22-4.50) |
| Educational level: High<sup>f</sup> | 1.72<br>(0.71-4.17) | 1.16<br>(0.45-2.99) | 2.07<br>(1.10-3.89) | 1.85<br>(0.95-3.59) | 0.46<br>(0.30-0.70) | 0.66<br>(0.41-1.04) |
| Sex: Male | 0.92<br>(0.42-1.98) | 1.01<br>(0.45-2.24) | 0.92<br>(0.51-1.65) | 0.93<br>(0.48-1.8) | 0.95<br>(0.67-1.36) | 1.05<br>(0.70-1.56) |
| Age | 0.97<br>(0.95-1.01) | 0.98<br>(0.95-1.01) | 1.01<br>(0.98-1.03) | 1.01<br>(0.99-1.03) | 0.98<br>(0.96-0.99) | 0.98<br>(0.96-0.99) |
| Travels to high-risk areas: Yes<sup>g</sup> | 2.41<br>(0.99-5.90) | 2.29<br>(0.90-5.78) | 2.03<br>(1.11-3.69) | 1.92<br>(1.04-3.52) | 0.54<br>(0.32-0.92) | 0.75<br>(0.43-1.32) |
| Work with patients or human tissues: Yes<sup>h</sup> | 0.88<br>(0.38-2.06) | 0.99<br>(0.40-2.43) | 0.70<br>(0.37-1.32) | 0.72<br>(0.37-1.4) | 0.54<br>(0.32-0.93) | 0.59<br>(0.32-1.07) |
| Hospital visits as a patient: Yes | 0.97<br>(0.34-2.79) | 1.00<br>(0.33-2.99) | 0.39<br>(0.05-2.91) | 0.42<br>(0.05-3.31) | 1.18<br>(0.65-2.16) | 1.02<br>(0.52-2.03) |
| Hospital visits as a professional: Yes | 0.48<br>(0.06-3.63) | 0.44<br>(0.05-3.50) | 1.28<br>(0.16-10.04) | 1.31<br>(0.15-11.23) | 0.59<br>(0.13-2.62) | 1.20<br>(0.22-6.46) |
| Use of antibiotics: Yes | 1.19<br>(0.52-2.72) | 1.09<br>(0.46-2.55) | 0.80<br>(0.35-1.82) | 0.86<br>(0.36-2.02) | 0.98<br>(0.64-1.49) | 1.28<br>(0.77-2.12) |
| Farm visits: Yes | 0.86<br>(0.32-2.34) | 0.99<br>(0.35-2.83) | 1.13<br>(0.43-2.95) | 1.34<br>(0.5-3.56) | 0.00<br>(0-Inf)<sup>i</sup> | 0.00<br>(0-Inf)<sup>i</sup> |

Notes:
<sup>a</sup>cOR: crude odds ratio.
<sup>b</sup>aOR: adjusted odds ratio.
<sup>c</sup>Nearby residents live within a 300 m radius from a WWTP.
<sup>d</sup>Data on Nearby residents in the Netherlands was not collected.
<sup>e</sup>Not possible to estimate the OR for WWTP workers because all workers in Germany had a negative stool sample result for ESBL-producing *E. coli*.
<sup>f</sup>Educational level according to the International Standard Classification of Education (ISCED): Low = ISCED 0-2 (Pre-primary education to Lower secondary education), High = ISCED ≥3 (Upper secondary education to Doctoral or equivalent).
<sup>g</sup>Travel to high risk areas for AR in the past year: Includes travels to North Africa, Sub-Saharan Africa, Asia, Central and South America, as well as the European countries Italy, Greece, Bulgaria and Slovenia.
<sup>h</sup>Work with patients or human tissues in the past year: Includes self-reported contact with patients at work and with human tissues (e.g. blood, urine, sputum, feces, vomit, saliva, or primary cell lines).
<sup>i</sup>Not possible to estimate the OR for farm visits because all participants who stated visiting a farm in the past year had a negative stool sample result for ESBL-producing *E. coli*.
ESBL: Extended-Spectrum Beta-Lactamases.
AR: Antibiotic Resistance.

Additionally, and according to our weighted and adjusted model, participants who reported traveling to high risk areas for AR in the past 12 months were almost twice as likely to have a positive result for ESBL-EC in stool samples, as compared to participants who did not travel to these high-risk areas (aOR 2.06, 95% CI: 1.33-3.19). None of the other covariates showed a statistically significant effect (see Online Resource Table 2 and Online Resource Fig. 1). The magnitude and direction of these estimates, as well as their confidence intervals, were fairly conserved in the stratified models by participation group (see Online Resource Fig. 2).

### Missing values

The highest proportion of missing values was found in the carriage of ESBL-EC (n = 163, 8.4%), driven mostly by the German population (n = 114, 24%, Table 1). A comparison of crude and adjusted odds ratios (OR) along with 95% CI for logistic regression models with complete case analysis and with the imputed dataset showed that the direction of effect did not change after imputation (Online Resource Table 3).

## Discussion

Across the three countries, we found no evidence of an increased risk for carriage of ESBL-EC neither in WWTP workers nor in residents living in close proximity to these WWTPs, as compared to the general population. We did find, however, evidence of increased odds for carriage of ESBL-EC in WWTP workers and in nearby residents in the Romanian population. Contrary to what we initially hypothesized, the effect for nearby residents was higher than the effect for WWTP workers in Romania.

An increased background prevalence of ESBL-EC in Romania, supported by our data, could be a risk factor for ESBL-EC carriage that sets the Romanian study population apart from the German and the Dutch. Additionally, travel to high-risk areas for AR has been identified as a risk factor for the carriage of ESBL-producing Enterobacterales because of the increased background prevalence of AR in some travel destinations [61–65]. Our data show that participants travel differently to high-risk areas for AR depending on their original country of residence. In Germany, our data collection took place in the south of the country where residents tend to choose Italy or Slovenia for their vacations because of the close geographical proximity, resulting in approximately half of the German participants reporting travels to high-risk areas for AR (Table 1).

Finding a higher ESBL-EC estimate for nearby residents than for WWTP workers in Romania, even after adjustment for other potential confounders and sources of exposure, suggests that the main source of exposure for nearby residents might not be the local WWTP. Potential sources of exposure for which we did not collect data and that might uniquely affect nearby residents in Romania but not WWTP workers are mentioned as follows. Risk factors for acquiring community-associated ESBL infection include use of corticosteroids [66] and personal history of diabetes mellitus [66,67], which is relevant for our study because, at 11.6%, Romania is one of the countries with the highest prevalence of diabetes mellitus in Europe [68]. Person-to-person transmission of ESBL-producing Enterobacterales within households has been documented in Spain [69], the Netherlands [70], and the U.S. [71], even showing identical strains between patients who had community-acquired infections with ESBL-producing Enterobacterales and their household members [72]. Additionally, ethnicity encodes cultural, social, and health behaviors that could result in a higher carriage rate for ESBL-EC [73]. From the door-to-door visits, differences in household size, sociodemographic characteristics, and underlying comorbidities were observed for nearby residents in Romania, although not systematically recorded. Therefore, these risk factors might differ between exposure groups in Romania at a greater degree than in the other countries.

Within the Romanian population, there is also a striking difference in travels to high-risk areas for AR depending on their participation group: although the proportion of participants among WWTP workers and the distant residents is similar regarding travels to high-risk areas for AR (30% and 33%), the proportion of nearby residents traveling to these high-risk areas for AR was, in comparison, low (10%). We observed a similar trend regarding educational level, where the proportion of highly educated participants in Romania was higher for WWTP workers and distant residents (97% and 91%) than for nearby residents (65%). In fact, when considering country of residence as an interaction term for the effect of participation group on carriage of ESBL-EC (Online Resource Table 4), the effect of Romania as country of residence alone disappeared (aOR 1.55, 95% CI: 0.79-3.05), while the effect of being a nearby resident in Romania carried the observed effect (aOR 5.49, 95% CI: 1.79-16.8). As frequency of travels and educational levels are proxies for socio-economic status (SES), we suspect that nearby residents in Romania have a lower SES, which would then affect our exposure-outcome relation. Although we did not directly collect data about SES, the constructed DAG (Fig. 2) confirmed that adjusting for other potential confounders is enough to find an unbiased estimate for the direct causal effect of proximity to WWTP (defined by participation group) on carriage of ESBL-EC. In our study, we did not measure the full extent of SES (only partially by e.g. education). Thus, SES is an unobserved confounder of the causal effect of participation group on carriage of ESBL-EC. It was therefore not possible to calculate an unbiased total effect of the exposure-outcome relation. However, adjusting for age, sex, education, country, travels to high risk areas for AR, antibiotics use, farm visits, work with patients or tissues, hospital visits as patients, and hospital visits as a professional made it possible to estimate the direct causal effect.

### Strengths and limitations

As far as we know, and despite the abundance of studies analyzing ARB in water and air samples from WWTPs [21–24], this is the first study investigating the carriage of ESBL-producing Enterobacterales in humans hypothesized to be exposed through ingestion of droplets, hand-to-mouth contact, or inhalation of aerosols due to close proximity to a WWTP, either from working at a WWTP or from living in the surroundings. Several characteristics make the AWARE study unique in its design. Data collection was conducted in three European countries with different background prevalences for AR. We explored the exposure-outcome relation defining two exposed groups and one comparison group, we followed a systematic sampling of participants adapted to the local regulations and logistical capabilities, we used reminders and incentives to increase participation, we developed our study questionnaire within a multidisciplinary team of experts, we used validated questions whenever possible, we conducted a pilot study to assess the feasibility of our methods, we conducted quality control processes for data input and data cleaning processes, we used standardized operating procedures (SOPs) in all three locations to guarantee laboratory methods to be comparable, and used positive controls for culture analyses. Additionally, we avoided using data-driven methods for variable selection. Instead, we conducted a thoughtful identification of potential confounders *a priori* with the help of a directed acyclic graph, and we used methods such as multiple imputation and inverse probability of sampling weights to analytically reduce the impact of missing values and low response. Our results are consistent in sensitivity analyses using alternative analytical methods to model our exposure-outcome relation: Traditional unweighted logistic regression models with complete case analysis and imputed analysis (Online Resource Table 3), unweighted stratified models by country (Table 2), model using country of residence as an interaction term (Online Resource Table 4).

Our study is, however, not exempt of limitations. Threats to internal validity include the risk of selection bias evidenced by the low participation response, especially in Germany and the Netherlands, for which we decided to use inverse probability of sampling weights. In our study, we suspect that the reasons for the observed low response in WWTP workers, nearby, and distant residents from Germany (response 22%, 6.95%, and 11%) and in the Netherlands (response 26%, and 6.4%) when compared with the response in Romania (response 62%, 53%, and 54%), reflect our recruitment methods and possibly background potential cultural differences among the countries. In Germany and in the Netherlands we invited potential participants using invitation letters sent by postal service, whereas in Romania we used a door-to-door approach because, in our experience, this method is more effective in Romania than postal letters. Also, studies involving stool samples have been reported to have a low response because of inherent reasons related to the nature of the stool sample [74,75]. These reasons put our study at risk of selection bias. Inverse probability of sampling weights has been described as an analytical method to adjust for selection bias where weights are assigned based on the factors that generate selection, which in our case is the response, and thus serve to reduce the differences between the study population and the target population [51,76].

Additionally, after recruitment and applying exclusion criteria for the analysis, we failed to reach the desired sample size for nearby residents in Germany and in Romania. We also failed to reach the desired sample size for workers in Germany at the recruitment stage. This has implications for the statistical power of our study to detect a desired effect, if there is in fact one. A *post hoc* power test restricted to study participants who completed all study phases (including providing a stool sample) shows that our data provides us with 63% and 75% statistical power to detect a minimum OR of 1.7 in workers and in nearby residents, when compared with distant residents.

Further, our data showed a proportion of 8% of missing values on the ESBL-EC carriage across countries (n = 163). Some of these missing values came from samples collected in the Netherlands (n = 4) and in Romania (n = 45) but the majority of the missing values for stool samples came from Germany (n = 114). Our data collection methods in Germany shed some light into this large number of missing values: only participants who had already completed the baseline questionnaire received a stool sample kit, and then were given a short time frame to hand in stool samples in person at the previously arranged time and place. These constrains were caused by the limited availability of the local microbiological laboratory to process samples, by the fact that we could not guarantee adequate preservation of samples if sent to the laboratory by postal service, and thus having to collect stool samples in person. Consequently, these values are missing completely at random or, worst case scenario, missing at random conditional on the country of residence. We are confident that randomness is key in the missing mechanism because participants would not have been able to self-assess their AR carriage status *a priori*. Besides fulfilling the randomness assumption for applying multiple imputation in our data, we performed *post hoc* imputation diagnostics by comparing models with complete cases vs. after imputation and did not find major differences in the directionality of estimates (Online Resource Table 3).

Finally, we have not included information about the heterogeneity of treatment processes in WWTPs across the three countries, nor have we included specific working conditions at the WWTP for the workers. Actual contact with raw wastewater can be limited to occasional sampling but could pose a higher threat of exposure depending on the time spent at certain locations within the WWTP, the type of activity performed, and the frequency of given activity, which are relevant factors for exposure intensity. Upcoming analyses from our project will include a formal exposure assessment for these study populations based on spatial techniques including physical distance of participants to the WWTPs, working conditions and preventive behavior at work for WWTP workers, and the specific operative characteristics of enrolled WWTPs.

## Conclusions

To the best of our knowledge, this is the first study investigating the carriage of ESBL-producing Enterobacterales in humans exposed to antibiotic resistant factors due to close proximity to a WWTP, either from working at a WWTP or from living in the surroundings. Using data collected in Germany, the Netherlands, and Romania, we did not find evidence of an increased risk of carriage of ESBL-producing *E. coli* in WWTP workers or in nearby residents across the three countries, as compared to the general population. We did find an increased risk for carriage of ESBL-EC in the subset of the Romanian population, both in WWTP workers and in nearby residents, which could be at least partially attributed to the local WWTP. However, this effect was higher for nearby residents than for workers, which suggests that, for nearby residents, unmeasured confounding factors could contribute to the increased carriage. Upcoming analyses from this project will perform exposure assessment using spatial techniques, including working conditions at WWTPs and working behavior from WWTP workers, and considering the heterogeneity of WWTP characteristics in terms of treatment efficacy and its consequences for the environment.

## Supporting information

Online Resource Fig. 1

Online Resource Fig. 2

Online Resource Table 1

Online Resource Table 2

Online Resource Table 3

Online Resource Table 4

## Data Availability

Data and materials are available upon request.

## Acknowledgements

We would like to thank Dr. Jana Bader at the Max von Pettenkofer Institute of LMU Munich for their support and expertise regarding analyses in Germany. We would also like to thank Nicole Stasch, Pezi Mang, Kim Weiszhar, Sonja Strieker, Marieke Behlen, and Nicole Schäfer for their hard work and support during the field phase of the study. Additionally, we extend our gratitude to WWTP workers, operators, nearby and distant residents around WWTP for their support, collaboration, and assistance during the sampling campaign and data collection. AWARE (Antibiotic Resistance in Wastewater: Transmission Risks for Employees and Residents around Wastewater Treatment Plants) is supported by the European Commission (JPI-EC-AMR ERA-Net Cofund grant no 681055), the Bundesministerum für Bildung und Forschung, DLR Projektträger (01KI1708), UEFISCDI project ERANET-JPI-EC-AMR-AWARE-WWTP No. 26/2017, the Netherlands Organisation for Health Research and Development, The Hague, the Netherlands (ZonMw, grant 547001007), and the Swedish Research Council VR Grant No. 2016-06512.

## Declarations

### Funding

AWARE (Antibiotic Resistance in Wastewater: Transmission Risks for Employees and Residents around Wastewater Treatment Plants) is supported by the European Commission (JPI-EC-AMR ERA-Net Cofund grant no 681055), the Bundesministerum für Bildung und Forschung, DLR Projektträger (01KI1708), UEFISCDI project ERANET-JPI-EC-AMR-AWARE-WWTP No. 26/2017, the Netherlands Organisation for Health Research and Development, The Hague, the Netherlands (ZonMw, grant 547001007, https://www.zonmw.nl/), and the Swedish Research Council VR Grant No. 2016-06512, all within the 5^th^ JPI AMR framework on transmission dynamics.

### Conflict of Interest

The authors have no conflicts of interest to declare that are relevant to the content of this article.

### Availability of data and material

Data and materials are available upon request.

### Code availability

Code is available upon request.

### Authors’ contributions

Study conception and design: MCC, DGJL, KR, DN, AW, AMRH, HS. Fieldwork and data collection: DRM, HB, MB, MK, LM, GP, LW. Microbiology: HB, MB, MK, LM, GP, MCC, BS, AW, HS. Data cleaning and analysis: DRM, HB, MB. Interpretation of the data: DRM, FB, HB, CFF, TW, LW, DGJL, KR, HS. Drafting of the manuscript: DRM. All authors read and approved the final manuscript.

### Ethics approval

This study was approved by the Ethics Committee of the University of Munich (LMU) (Project-No. 17-734) and by the Research Ethics Committee of the University of Bucharest (Registration-No. 164/05.12.2017). In the Netherlands, this research is exempted for ethical approval under the Dutch Medical Research Involving Human Subjects Act (WMO; Committee: Medisch Ethische Toetsingscommissie, number of confirmation: 19-001/C). All procedures performed in studies involving human participants were in accordance with the ethical standards of the institutional and national research committees and with the 1964 Helsinki declaration and its later amendments or comparable ethical standards, as well as with Directive 95/46/EC, and the 1977 Oviedo Convention of the Council of Europe on human rights and biomedicine.

### Consent to participate

Written informed consent was obtained from all individual participants included in the study and their legal guardians when applicable.

## Captions

- Online Resource Table 1: Descriptive characteristics of ESBL-producing *E. coli* carriers by country, n = 1940, AWARE Study, 2021
- Online Resource Table 2: Comparison of models estimating the effect of participation group (wastewater treatment plant -WWTP-worker, nearby resident, distant resident) as a proxy for exposure routes (ingestion of droplets, hand-to-mouth contact, or inhalation of aerosols) in and around the local WWTP on the presence of ESBL-producing E. coli in stool samples, AWARE Study, 2021
- Online Resource Table 3: Multiple imputation diagnostics - Traditional (unweighted) logistic regression models, complete cases vs. imputed, AWARE Study, 2021
- Online Resource Table 4: Interaction model with an interaction term for participation group with country for the carriage of ESBL-producing E. coli in comparison to crude estimates from a traditional unweighted logistic regression model, n = 1940, AWARE Study, 2021
- Online Resource Fig. 1: Comparison of models estimating the effect of participation group (wastewater treatment plant - WWTP-worker, nearby resident, distant resident) as a proxy for exposure routes (ingestion of droplets, hand-to-mouth contact, or inhalation of aerosols) in and around the local WWTP on the presence of ESBL-producing E. coli in stool samples (all estimates are shown), AWARE Study, 2021. Models adjusted for age, sex, education, country, travels to high risk areas,working with human tissues, antibiotic use, farm visits, hospital visits as patient and hospital visits as a professional. IPW: Inverse Probability Weighted model. ref. = Reference level. Travel to high risk areas for AR in the past year includes travels to North Africa, Sub-Saharan Africa, Asia, Central and South America, as well as the European countries Italy, Greece, Bulgaria and Slovenia. Crude: Model with only the given variable, ignoring potential covariates. Adjusted: Model with the given variable, including all potential covariates in the exposure-outcome relation. Unweighted: Model without applying inverse probability weights (IPW). Weighted: Model applying inverse probability weights (IPW). See text for details
- Online Resource Fig. 2: Comparison of models estimating the effect of participation group (wastewater treatment plant - WWTP-worker, nearby resident, distant resident) as a proxy for exposure routes (ingestion of droplets, hand-to-mouth contact, or inhalation of aerosols) in and around the local WWTP on the presence of ESBL-producing E. coli in stool samples, stratified by participation group, AWARE Study, 2021. Models adjusted for age, sex, education, country, travels to high risk areas, working with human tissues, antibiotic use, farm visits, hospital visits as patient and hospital visits as a professional. IPW: Inverse Probability Weighted model. ref. = Reference level. Travel to high risk areas for AR in the past year includes travels to North Africa, Sub-Saharan Africa, Asia, Central and South America, as well as the European countries Italy, Greece, Bulgaria and Slovenia. Crude: Model with only the given variable, ignoring potential covariates. Adjusted: Model with the given variable, including all potential covariates in the exposure-outcome relation. Unweighted: Model without applying inverse probability weights (IPW). Weighted: Model applying inverse probability weights (IPW). See text for details

## Notes

### Competing Interest Statement

The authors have declared no competing interest.

### Clinical Protocols

https://www.mdpi.com/2079-6382/10/5/478

### Author Declarations

Ethics approval This study was approved by the Ethics Committee of the University of Munich (LMU) (Project-No. 17- 734) and by the Research Ethics Committee of the University of Bucharest (Registration-No. 164/05.12.2017). In the Netherlands, this research is exempted for ethical approval under the Dutch Medical Research Involving Human Subjects Act (WMO; Committee: Medisch Ethische Toetsingscommissie, number of confirmation: 19-001/C). All procedures performed in studies involving human participants were in accordance with the ethical standards of the institutional and national research committees and with the 1964 Helsinki declaration and its later amendments or comparable ethical standards, as well as with Directive 95/46/EC, and the 1977 Oviedo Convention of the Council of Europe on human rights and biomedicine. Consent to participate Written informed consent was obtained from all individual participants included in the study and their legal guardians when applicable.

