## Supplementary material for "Carriage of ESBL-producing Enterobacterales in wastewater treatment plant workers and surrounding residents - The AWARE Study": Online Resource Fig. 1

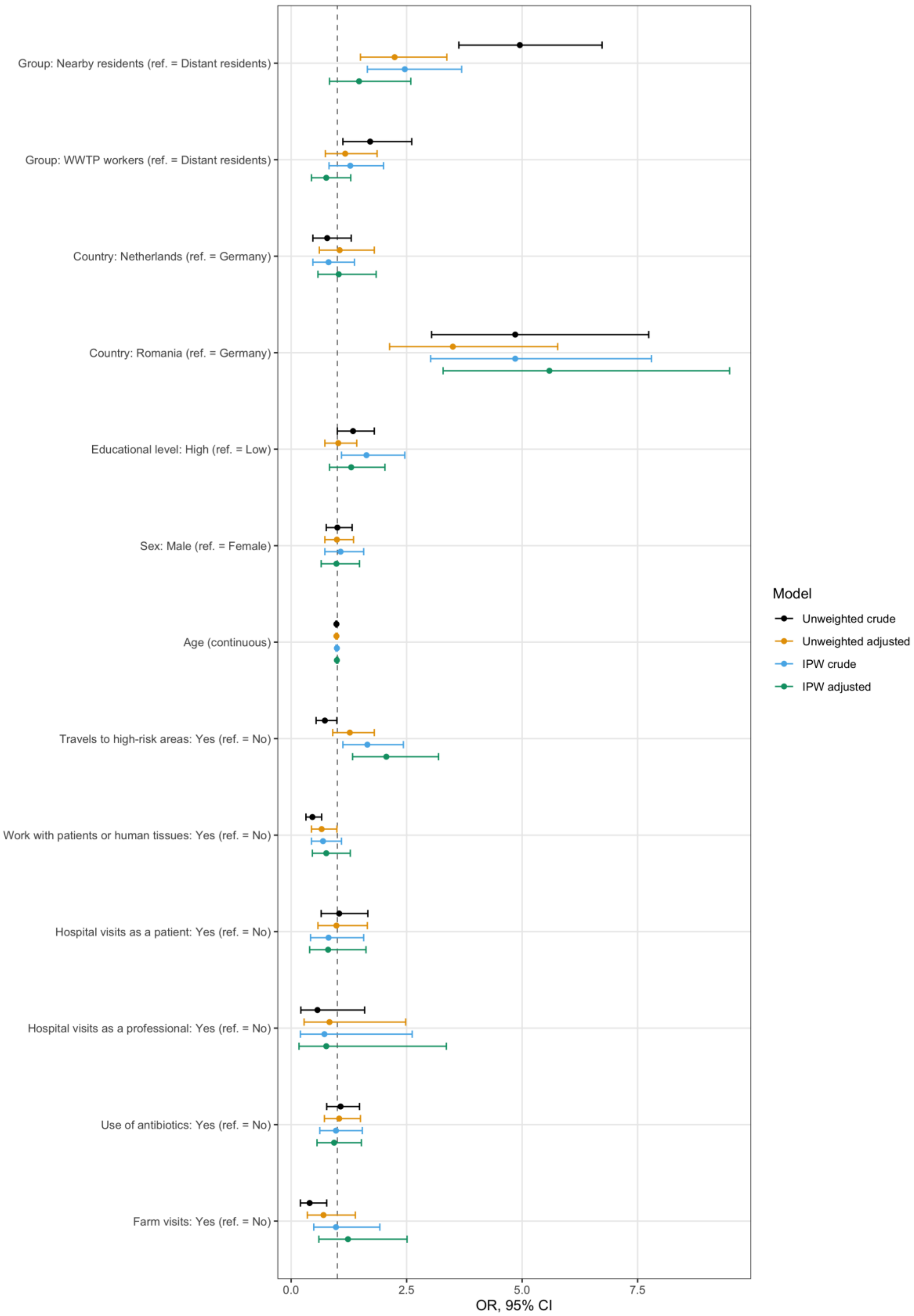


Online Resource Fig. 1: Comparison of models estimating the effect of participation group (wastewater treatment plant -WWTP- worker, nearby resident, distant resident) as a proxy for exposure routes (ingestion of droplets, hand-to-mouth contact, or inhalation of aerosols) in and around the local WWTP on the presence of ESBL-producing E. coli in stool samples (all estimates are shown), AWARE Study, 2021.
Models adjusted for age, sex, education, country, travels to high risk areas, working with human tissues, antibiotic use, farm visits, hospital visits as patient and hospital visits as a professional.
IPW: Inverse Probability Weighted model.

ref. = Reference level.
Travel to high risk areas for AR in the past year includes travels to North Africa, Sub-Saharan Africa, Asia, Central and South America, as well as the European countries Italy, Greece, Bulgaria and Slovenia.
Crude: Model with only the given variable, ignoring potential covariates.
Adjusted: Model with the given variable, including all potential covariates in the exposure-outcome relation.
Unweighted: Model without applying inverse probability weights (IPW).
Weighted: Model applying inverse probability weights (IPW).
See text for details

### Supplementary Information

#### Carriage of ESBL-producing Enterobacterales in wastewater treatment plant workers and surrounding residents - The AWARE Study

Daloha Rodríguez-Molina[1,2,3], Fanny Berglund[4,5], Hetty Blaak[6], Marcela Popa[7,8], Carl-Fredrik Flach[4,5], Merel Kemper[6], Luminita Marutescu[7,8], Gratiela Pircalabioru[7,8], Beate Spießberger[9,10,11], Tobias Weinmann[1], Laura Wengenroth[1], Mariana Carmen Chifiriuc[7,8], D. G. Joakim Larsson[4,5], Katja Radon[1], Dennis Nowak[1,12], Andreas Wieser[9,10,11], Ana Maria de Roda Husman[6], Heike Schmitt[6].

1: Occupational and Environmental Epidemiology & NetTeaching Unit; Institute and Clinic for Occupational, Social and Environmental Medicine; University Hospital, LMU Munich, Munich, Germany.

2: Institute for Medical Information Processing, Biometry, and Epidemiology – IBE, LMU Munich, Munich, Germany.

3: Pettenkofer School of Public Health, Munich, Germany.

4: Department of Infectious Diseases, Institute of Biomedicine, The Sahlgrenska Academy, University of Gothenburg, Gothenburg, Sweden.

5: Centre for Antibiotic Resistance Research (CARe), University of Gothenburg, Gothenburg, Sweden.

6: Centre of Infectious Disease Control, National Institute for Public Health and the Environment, Bilthoven, The Netherlands.

7: Department of Microbiology and Immunology, Faculty of Biology, University of Bucharest and the Academy of Romanian Scientists, Bucharest, Romania.

8: Earth, Environmental and Life Sciences Section, Research Institute of the University of Bucharest, University of Bucharest, Bucharest, Romania.

9: Department of Infectious Diseases and Tropical Medicine, LMU University Hospital Munich, Munich, Germany.

10: German Centre for Infection Research (DZIF) Partner Site Munich.

11: Max von Pettenkofer Institute, Faculty of Medicine, LMU Munich.

12: Comprehensive Pneumology Center Munich (CPC-M), German Center for Lung Research (DZL), Munich, Germany.

**Corresponding author**: Daloha Rodríguez-Molina; Occupational and Environmental Epidemiology and & NetTeaching Unit; Institute and Clinic for Occupational, Social and Environmental Medicine;

University Hospital, LMU Munich; Ziemssenstr. 1, D-80336 Munich, Germany; Tel.: +49 (89) 4400-52358 Fax.: +49 (89) 4400-54954;
