## Supplementary material for "Carriage of ESBL-producing Enterobacterales in wastewater treatment plant workers and surrounding residents - The AWARE Study": Online Resource Fig. 2

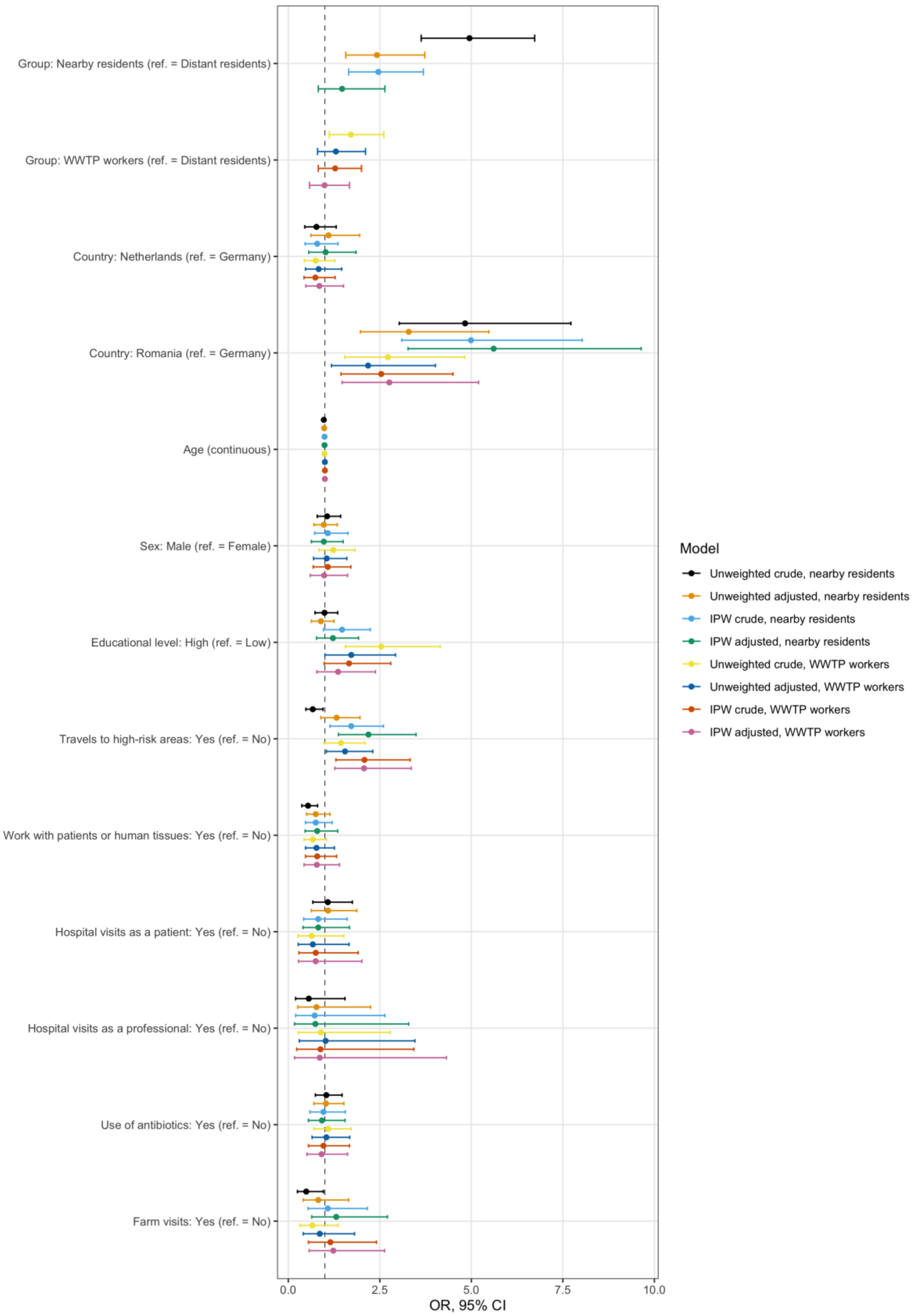


Online Resource Fig. 2: Comparison of models estimating the effect of participation group (wastewater treatment plant -WWTP- worker, nearby resident, distant resident) as a proxy for exposure routes (ingestion of droplets, hand-to-mouth contact, or inhalation of aerosols) in and around the local WWTP on the presence of ESBL-producing E. coli in stool samples, stratified by participation group, AWARE Study, 2021.
