## Supplementary material for "Carriage of ESBL-producing Enterobacterales in wastewater treatment plant workers and surrounding residents - The AWARE Study": Online Resource Table 1

*Online Resource Table 1: Descriptive characteristics of ESBL-producing E. coli carriers by country, n = 1940, AWARE Study, 2021*

| | | | Overall | | Germany | | The Netherlands | | Romania | |
|---|---|---|---|---|---|---|---|---|---|---|
| **Variable** | **Missings** | **level** | **Positive** | **p** | **Positive** | **p** | **Positive** | **p** | **Positive** | **p** |
| n | | | 236 | | 26 | | 47 | | 163 | |
| Age, years (median [IQR]) | 0 | | 43 [31, 55] | <0.001 | 38 [31, 51] | 0.051 | 55 [48, 60] | 0.977 | 40 [29, 52] | 0.005 |
| Sex, n (%) | 4 | Female | 122 (13) | 0.959 | 15 (8) | 0.905 | 25 (6) | 0.878 | 82 (28) | 0.919 |
| | | Male | 114 (13) | | 11 (7) | | 22 (5) | | 81 (27) | |
| Highest educational level obtained, n (%)[a] | 8 | Low | 75 (11) | 0.068 | 6 (4) | 0.189 | 15 (4) | 0.033 | 54 (41) | <0.001 |
| | | High | 161 (14) | | 20 (9) | | 32 (8) | | 109 (24) | |
| Participation group, n (%) | 0 | Distant resident[b] | 79 (7) | <0.001 | 21 (8) | 0.259 | 40 (4) | 0.569 | 18 (12) | <0.001 |
| | | Nearby resident[c] | 123 (29) | | 5 (6) | | --- (---)[d] | | 118 (36) | |
| | | WWTP worker | 34 (11) | | 0 (0) | | 7 (6) | | 27 (23) | |
| Work with patients or human tissues in the past year, n (%)[e] | 43 | No | 186 (16) | <0.001 | 15 (7) | 1.000 | 32 (7) | 0.333 | 139 (29) | 0.042 |
| | | Yes | 43 (8) | | 9 (7) | | 15 (5) | | 19 (19) | |
| Hospital visits as a patient in the past year, n (%) | 2 | No | 215 (13) | 0.889 | 22 (7) | 1.000 | 46 (6) | 0.505 | 147 (27) | 0.634 |
| | | Yes | 21 (14) | | 4 (7) | | 1 (2) | | 16 (30) | |
| Hospital visits as a professional in the past year, n (%) | 2 | No | 232 (13) | 0.395 | 25 (7) | 1.000 | 46 (6) | 0.565 | 161 (28) | 0.734 |
| | | Yes | 4 (8) | | 1 (4) | | 1 (7) | | 2 (20) | |
| Use of antibiotics in the past year, n (%) | 4 | No | 177 (13) | 0.703 | 17 (7) | 0.810 | 40 (6) | 0.739 | 120 (27) | 1.000 |
| | | Yes | 56 (14) | | 9 (8) | | 7 (5) | | 40 (27) | |
| Farm visits in the past year, n (%)[f] | 9 | No | 226 (14) | 0.005 | 22 (7) | 1.000 | 42 (6) | 0.799 | 162 (28) | 0.015 |
| | | Yes | 9 (6) | | 4 (6) | | 5 (6) | | 0 (0) | |
| Travel to high risk areas for AR in the past year, n (%) | 18 | No | 169 (14) | 0.031 | 6 (3) | 0.019 | 22 (4) | 0.023 | 141 (30) | 0.010 |
| | | Yes | 62 (11) | | 19 (10) | | 24 (8) | | 19 (17) | |

Notes:
[a]Educational level according to the International Standard Classification of Education (ISCED): Low = ISCED 0-2 (Pre-primary education to Lower secondary education), High = ISCED ≥3 (Upper secondary education to Doctoral or equivalent).
[b]Distant residents live at least 1000 m away from a WWTP
[c]Nearby residents live within a 300 m radius from a WWTP.
[d]No data from nearby residents were collected in the Netherlands.
[e]Work with patients or human tissues in the past year: Includes self-reported contact with patients at work and with human tissues (e.g. blood, urine, sputum, feces, vomit, saliva, or primary cell lines).
[f]Travel to high risk areas for AR in the past year: Includes travels to North Africa, Sub-Saharan Africa, Asia, Central and South America, as well as the European countries Italy, Greece, Bulgaria and Slovenia.
ESBL: Extended-Spectrum Beta-Lactamases; AR: Antibiotic Resistance.
