## Supplementary material for "Carriage of ESBL-producing Enterobacterales in wastewater treatment plant workers and surrounding residents - The AWARE Study": Online Resource Table 2

*Online Resource Table 2: Comparison of models estimating the effect of participation group (wastewater treatment plant -WWTP- worker, nearby resident, distant resident) as a proxy for exposure routes (ingestion of droplets, hand-to-mouth contact, or inhalation of aerosols) in and around the local WWTP on the presence of ESBL-producing E. coli in stool samples, AWARE Study, 2021*

| | **Unweighted cOR (95% CI)[a]** | **Unweighted aOR (95% CI)[b]** | **IPW cOR (95% CI)[a,c]** | **IPW aOR (95% CI)[b,c]** |
|---|---|---|---|---|
| Group: Nearby resident [d] | 4.95 (3.63-6.73) | 2.24 (1.5-3.37) | 2.46 (1.65-3.69) | 1.47 (0.83-2.59) |
| Group: WWTP worker | 1.71 (1.12-2.61) | 1.17 (0.74-1.86) | 1.28 (0.82-2) | 0.76 (0.44-1.29) |
| Country: Netherlands | 0.78 (0.47-1.3) | 1.05 (0.61-1.8) | 0.81 (0.47-1.37) | 1.03 (0.58-1.84) |
| Country: Romania | 4.85 (3.04-7.74) | 3.5 (2.13-5.77) | 4.85 (3.02-7.8) | 5.59 (3.29-9.49) |
| Educational level: High[e] | 1.34 (1-1.8) | 1.02 (0.73-1.42) | 1.63 (1.09-2.46) | 1.3 (0.83-2.03) |
| Sex: Male | 1 (0.76-1.32) | 0.99 (0.73-1.35) | 1.07 (0.73-1.57) | 0.98 (0.65-1.48) |
| Age | 0.98 (0.97-0.99) | 0.98 (0.97-1) | 0.99 (0.97-1) | 0.99 (0.98-1.01) |
| Travels to high-risk areas: Yes[f] | 0.73 (0.54-0.99) | 1.27 (0.9-1.8) | 1.65 (1.12-2.43) | 2.06 (1.33-3.19) |
| Work with patients or human tissues: Yes[g] | 0.46 (0.32-0.66) | 0.66 (0.44-0.99) | 0.69 (0.44-1.09) | 0.76 (0.46-1.28) |
| Hospital visits as a patient: Yes | 1.04 (0.65-1.66) | 0.98 (0.58-1.65) | 0.81 (0.42-1.57) | 0.8 (0.4-1.62) |
| Hospital visits as a professional: Yes | 0.57 (0.21-1.59) | 0.83 (0.28-2.48) | 0.72 (0.2-2.62) | 0.76 (0.17-3.36) |
| Use of antibiotics: Yes | 1.07 (0.77-1.48) | 1.04 (0.72-1.5) | 0.97 (0.62-1.54) | 0.93 (0.56-1.52) |
| Farm visits: Yes | 0.4 (0.2-0.77) | 0.7 (0.35-1.39) | 0.97 (0.49-1.92) | 1.23 (0.6-2.51) |

Notes:
[a]cOR: crude odds ratio.
[b]aOR: adjusted odds ratio.
[,c]IPW: Inverse Probability Weighted model.
[d]Nearby residents live within a 300 m radius from a WWTP.
[e]Educational level according to the International Standard Classification of Education (ISCED): Low = ISCED 0-2 (Pre-primary education to Lower secondary education), High = ISCED ≥3 (Upper secondary education to Doctoral or equivalent).
[f]Travel to high risk areas for AR in the past year: Includes travels to North Africa, Sub-Saharan Africa, Asia, Central and South America, as well as the European countries Italy, Greece, Bulgaria and Slovenia.
[g]Work with patients or human tissues in the past year: Includes self-reported contact with patients at work and with human tissues (e.g. blood, urine, sputum, feces, vomit, saliva, or primary cell lines).
ESBL: Extended-Spectrum Beta-Lactamases.
AR: Antibiotic Resistance.
