## Supplementary material for "Carriage of ESBL-producing Enterobacterales in wastewater treatment plant workers and surrounding residents - The AWARE Study": Online Resource Table 3

*Online Resource Table 3: Multiple imputation diagnostics - Traditional (unweighted) logistic regression models, complete cases vs. imputed, AWARE Study, 2021*

| | | Crude models, OR (95% CI) | | Adjusted models, OR (95% CI) | |
|---|---|---|---|---|---|
| | **Missings** | **Complete cases** | **Imputed, n = 1940** | **Complete cases, n = 1707** | **Imputed, n = 1940** |
| Group: Nearby resident [d] | 163 | 5.16 (3.79-7.06) | 4.95 (3.63-6.73) | 2.86 (1.87-4.45) | 2.24 (1.5-3.37) |
| Group: WWTP worker | 163 | 1.56 (1.01-2.37) | 1.71 (1.12-2.61) | 1.09 (0.64-1.83) | 1.17 (0.74-1.86) |
| Country: Netherlands | 163 | 0.79 (0.49-1.32) | 0.78 (0.47-1.3) | 1.37 (0.79-2.45) | 1.05 (0.61-1.8) |
| Country: Romania | 163 | 5 (3.28-7.91) | 4.85 (3.04-7.74) | 3.57 (2.16-6.11) | 3.5 (2.13-5.77) |
| Age | 163 | 0.97 (0.96-0.98) | 0.98 (0.97-0.99) | 0.98 (0.97-0.99) | 0.98 (0.97-1) |
| Sex: Male | 167 | 0.98 (0.75-1.29) | 1 (0.76-1.32) | 1.01 (0.73-1.39) | 0.99 (0.73-1.35) |
| Educational level: High[e] | 167 | 1.33 (0.99-1.79) | 1.34 (1-1.8) | 1.04 (0.74-1.47) | 1.02 (0.73-1.42) |
| Travels to high-risk areas: Yes[f] | 180 | 0.7 (0.51-0.95) | 0.73 (0.54-0.99) | 1.34 (0.93-1.93) | 1.27 (0.9-1.8) |
| Work with patients or human tissues: Yes[g] | 198 | 0.45 (0.31-0.63) | 0.46 (0.32-0.66) | 0.69 (0.46-1.02) | 0.66 (0.44-0.99) |
| Hospital visits as a patient: Yes | 165 | 1.07 (0.64-1.69) | 1.04 (0.65-1.66) | 1.06 (0.6-1.82) | 0.98 (0.58-1.65) |
| Hospital visits as a professional: Yes | 165 | 0.56 (0.17-1.39) | 0.57 (0.21-1.59) | 0.83 (0.23-2.33) | 0.83 (0.28-2.48) |
| Use of antibiotics: Yes | 167 | 1.08 (0.78-1.48) | 1.07 (0.77-1.48) | 1.03 (0.7-1.5) | 1.04 (0.72-1.5) |
| Farm visits: Yes | 172 | 0.37 (0.17-0.69) | 0.4 (0.2-0.77) | 0.75 (0.34-1.46) | 0.7 (0.35-1.39) |
