## Supplementary material for "Carriage of ESBL-producing Enterobacterales in wastewater treatment plant workers and surrounding residents - The AWARE Study": Online Resource Table 4

*Online Resource Table 4: Interaction model with an interaction term for participation group with country for the carriage of ESBL-producing E. coli in comparison to crude estimates from a traditional unweighted logistic regression model, n = 1940, AWARE Study, 2021*

| | **cOR (95% CI)[a]** | **With interaction, aOR (95% CI)[b]** |
|---|---|---|
| Group: Nearby resident [c] | 4.95 (3.63-6.73) | 0.73 (0.27-1.95) |
| Group: WWTP worker | 1.71 (1.12-2.61) | 0 (0-Inf)[d] |
| Country: Netherlands | 0.78 (0.47-1.3) | 0.79 (0.45-1.39) |
| Country: Romania | 4.85 (3.04-7.74) | 1.55 (0.79-3.05) |
| Educational level: High[e] | 1.34 (1-1.8) | 0.98 (0.7-1.37) |
| Sex: Male | 1 (0.76-1.32) | 1.03 (0.76-1.41) |
| Age | 0.98 (0.97-0.99) | 0.98 (0.97-0.99) |
| Travels to high-risk areas: Yes[f] | 0.73 (0.54-0.99) | 1.31 (0.92-1.86) |
| Work with patients or human tissues: Yes[g] | 0.46 (0.32-0.66) | 0.69 (0.46-1.04) |
| Hospital visits as a patient: Yes | 1.04 (0.65-1.66) | 0.96 (0.57-1.62) |
| Hospital visits as a professional: Yes | 0.57 (0.21-1.59) | 0.83 (0.28-2.46) |
| Use of antibiotics: Yes | 1.07 (0.77-1.48) | 1.04 (0.72-1.5) |
| Farm visits: Yes | 0.4 (0.2-0.77) | 0.72 (0.36-1.45) |
| Interaction: NL x Nearby residents | --- | [h] |
| Interaction: NL x WWTP workers | --- | 881070.99 (0-Inf)[d] |
| Interaction: RO x Nearby residents | --- | 5.49 (1.79-16.8) |
| Interaction: RO x WWTP workers | --- | 2385014.82 (0-Inf)[d] |

Notes:
[a]cOR: crude odds ratio.
[b]aOR: adjusted odds ratio.
[c]Nearby residents live within a 300 m radius from a WWTP.
[d]Not possible to estimate the OR for WWTP workers because all workers in Germany had a negative stool sample result for ESBL-producing E. coli.
[e]Educational level according to the International Standard Classification of Education (ISCED): Low = ISCED 0-2 (Pre-primary education to Lower secondary education), High = ISCED ≥3 (Upper secondary education to Doctoral or equivalent).
[f]Travel to high risk areas for AR in the past year: Includes travels to North Africa, Sub-Saharan Africa, Asia, Central and South America, as well as the European countries Italy, Greece, Bulgaria and Slovenia.
[g]Work with patients or human tissues in the past year: Includes self-reported contact with patients at work and with human tissues (e.g. blood, urine, sputum, feces, vomit, saliva, or primary cell lines).
[h]Data on Nearby residents in the Netherlands was not collected. There is, therefore, no interaction term for NL x Nearby resident.
ESBL: Extended-Spectrum Beta-Lactamases.
AR: Antibiotic Resistance.
